# High-intensity exercise is a risk factor for renal medullary carcinoma in individuals with sickle cell trait

**DOI:** 10.1101/2021.02.23.21252313

**Authors:** Daniel D. Shapiro, Melinda Soeung, Luigi Perelli, Eleonora Dondossola, Devaki Shilpa Surasi, Durga N. Tripathi, Jean-Philippe Bertocchio, Ruohan Xia, Michael W. Starbuck, Michael L. Van Alstine, Priya Rao, Matthew H. G. Katz, Nathan H. Parker, Amishi Y. Shah, Alessandro Carugo, Timothy P. Heffernan, Keri L. Schadler, Christopher Logothetis, Cheryl L. Walker, Christopher G. Wood, Jose A. Karam, Giulio F. Draetta, Nizar M. Tannir, Giannicola Genovese, Pavlos Msaouel

**Affiliations:** Department of Urology, The University of Texas MD Anderson Cancer Center, Houston, TX 77030, USA; Department of Genomic Medicine, The University of Texas MD Anderson Cancer Center, Houston, TX 77030, USA; Department of Genitourinary Medical Oncology, The University of Texas MD Anderson Cancer Center, Houston, TX 77030, USA; David H. Koch Center for Applied Research of Genitourinary Cancers, The University of Texas, MD Anderson Cancer Center, Houston, TX 77030, USA; Department of Nuclear Imaging, The University of Texas MD Anderson Cancer Center, Houston, TX 77030, USA; Center for Precision Environmental Health, Baylor College of Medicine, Houston, TX 77030, USA; The Ronin Project, San Mateo, CA, USA; Department of Pathology, The University of Texas MD Anderson Cancer Center, Houston, TX 77030, USA; Department of Surgical Oncology, The University of Texas MD Anderson Cancer Center, Houston, TX 77030, USA; Department of Behavioral Science, The University of Texas MD Anderson Cancer Center, Houston, TX 77030, USA; Institute for Applied Cancer Science, The University of Texas MD Anderson Cancer Center, Houston, TX 77030, USA; Translational Research to Advance Therapeutics and Innovation in Oncology (TRACTION), The University of Texas MD Anderson Cancer Center, Houston, TX 77030, USA; Department of Pediatrics, The University of Texas MD Anderson Cancer Center, Houston, TX 77030, USA; Department of Translational Molecular Pathology, The University of Texas MD Anderson Cancer Center, Houston, TX 77030, USA

## Abstract

Renal medullary carcinoma (RMC) predominantly occurs in individuals with sickle cell trait (SCT). We found that patients with RMC more frequently participated in high-intensity exercise than matched controls, and renal medullary hypoxia significantly increased predominantly in the right kidney of mice with SCT following high-but not moderate-intensity exercise, consistent with the distinct predilection of RMC toward the right kidney. These results establish high-intensity exercise as a risk factor for RMC in individuals with SCT.

---

Renal medullary carcinoma (RMC) is a lethal cancer that arises from the renal medulla and has a median survival of only 13 months despite best available treatments.^1^ RMC almost exclusively occurs in the setting of hemoglobinopathies that cause red blood cell (RBC) sickling in the renal medulla, of which sickle cell trait (SCT) is the most common.^1–3^ Previous reports hypothesized that the pathogenesis of RMC may relate to tissue hypoxia caused by RBC sickling in the renal medulla, particularly in the right kidney due to the increased vascular resistance induced by the longer length of the right compared with the left renal artery.^1,2^ Additionally, high-intensity exercise among individuals with SCT increases the risk of rhabdomyolysis and sudden cardiac death secondary to tissue hypoxia from RBC sickling.^4,5^ To date, however, the renal sequela of exercise in the setting of SCT have not been investigated, and studies of RMC have not identified modifiable risk factors.

We utilized a large dataset of RMC patients and complementary animal models of SCT to investigate the relationship between high-intensity exercise, renal hypoxia, and RMC. To determine whether patients with RMC have higher rates of reported high-intensity exercise, we reviewed the records of patients with RMC diagnosed at our institution. We compared RMC patients (n=71) to an age- and biological sex–matched cohort of patients (n=122) also treated at our institution for other advanced genitourinary malignancies (**Supplementary Tables 1–4**). As is typical of the disease,^1–3^ all RMC patients had a sickle hemoglobinopathy (predominantly SCT), the majority were male (69%) and of African descent (86%), and the primary tumor was located in the right kidney three times more frequently (76%) than the left (**Supplementary Tables 1–2**). Compared with matched control patients, RMC patients had significantly higher rates of positive activity index, reported exercise, athletic involvement, and military service (*P* ≤ 0.005 for all) (**Supplementary Table 1**). Additionally, we used cross-sectional imaging obtained prior to systemic therapy initiation to digitally quantify the skeletal muscle (SM) surface area and found it to be significantly higher in RMC patients (median 59.3 cm^2^/m^2^, IQR 50.2-69.9 cm^2^/m^2^) compared with controls (median 52.4 cm^2^/m^2^, IQR 45.0-59.1 cm^2^/m^2^) (*P* = 0.001) (**Fig. 1a-b**). Causal models using directed acyclic graphs identified race, age, and biological sex as potential confounders for the associations between RMC diagnosis, exercise activity, and SM surface area (**Supplementary Fig. 1**). Multivariable models adjusting for these confounders demonstrated a significant relationship between physical activity index and RMC diagnosis (*P* < 0.001), as well as between higher SM surface area and RMC diagnosis (*P* = 0.03) (**Fig. 1c**).

**Figure 1.**
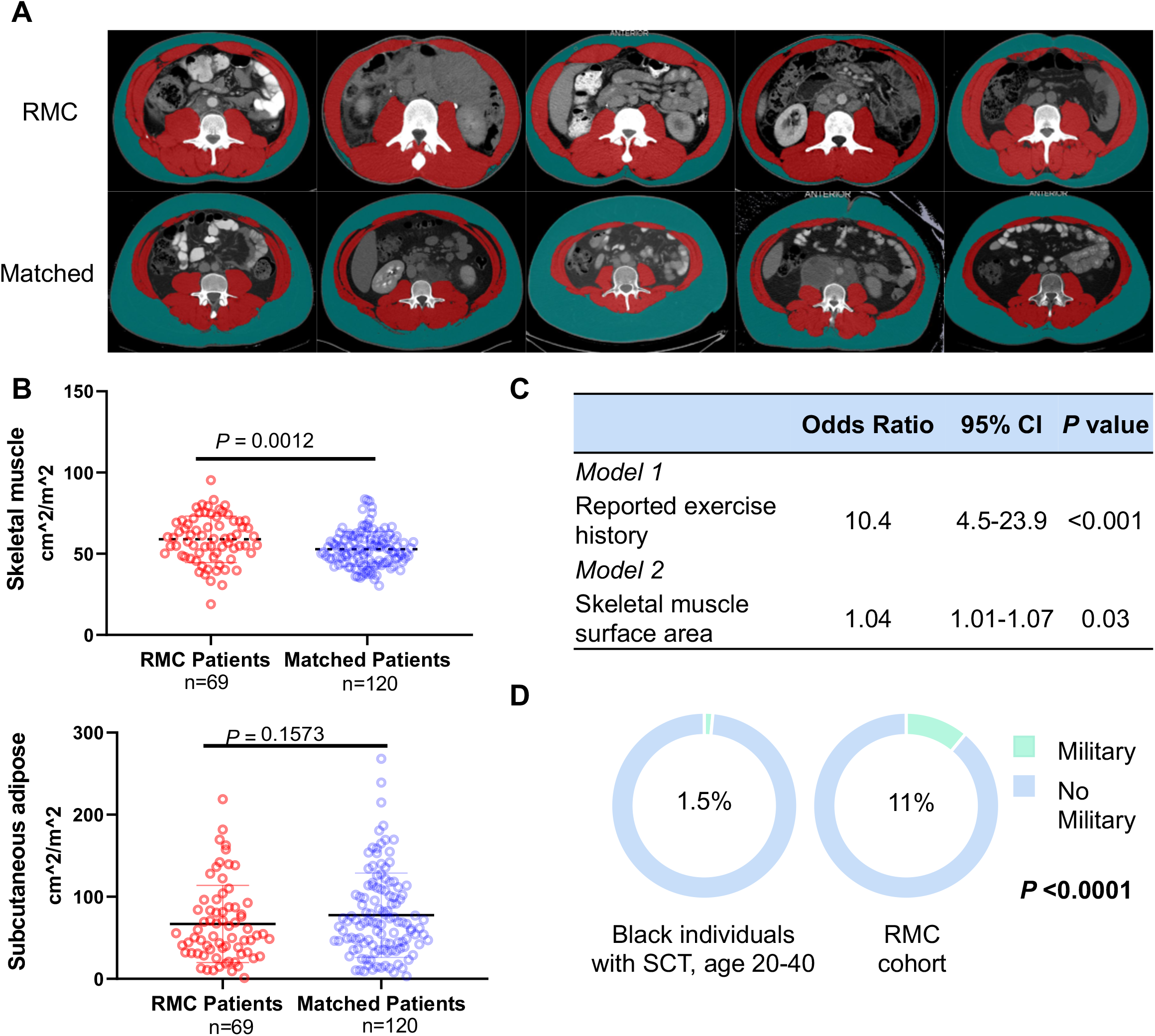
**(A)** Representative axial CT images from five RMC (top) and matched control (bottom) patients analyzed for skeletal muscle (SM) surface area (red) and subcutaneous adipose (green). **(B)** Dot plots of SM (top) and subcutaneous adipose (bottom) surface area. **(C)** Multivariable logistic regression models of the association between exercise history (model 1) or skeletal muscle surface area (model 2) and RMC. **(D)** Estimated proportion of black individuals with SCT aged 20 to 40 years in the United States military (1.5%) compared to the proportion of similarly aged black patients with SCT in the RMC cohort (11%).

We validated these findings in a separate epidemiologic comparison group by looking at the proportion of all black individuals with SCT who serve in the U.S. military, hypothesizing that the intense physical activity of military duty may increase the risk of RMC. We found that a higher proportion (OR 8.4, 95% CI 3.2-18.4, *P* < 0.0001) of patients with RMC have served in the military than would be expected compared to the similarly aged black population with SCT in the United States (**Fig. 1d**). We subsequently prospectively gathered detailed information on the exercise activity of 7 additional RMC patients seen at our institution (**Supplementary tables 5–6)**. Consistent with our retrospective findings, the majority of patients (71%) reported frequent, high-intensity exercise defined as ≥80% of maximal heart rate during each exercise session (**Supplementary table 6**).

To further interrogate our hypothesis, we generated a genetically engineered mouse model (GEMM) of SCT that leverages fluorescence for imaging purposes. The *CDH16*^*Cre*^ strain was crossed with the conditional *Rosa26*^*LSL-TdT*^ strain and the hα/hα::β^S^/β^S^ strain to generate a GEMM of SCT (hα/hα::β^A^/β^S^) that allows for tissue specific activation of TdTomato (TdT) fluorescence reporter in the kidney epithelium. (**Supplementary Fig. 2a**). Pathological evaluation confirmed RBC sickling in mice with SCT (**Supplementary Fig. 2c**). Using the contrast between TdT fluorescence reporter and injected dextran conjugated with fluorescein isothiocyanate (FITC), we evaluated the microvasculature integrity and flow in the renal inner medulla. Mice with SCT had discontinuous and disorganized blood vessels, including significantly shorter diameters (*P* < 0.0001) and lengths (*P* < 0.0001) of healthy blood vessels (**Fig. 2a-c**). To measure whether hypo-perfusion in blood vessels lead to hypoxia, we injected Hypoxyprobe, confirming that these microvascular changes yielded significantly higher hypoxia in the renal inner medulla of SCT mice compared with wild-type controls (*P* < 0.0001) (**Supplementary Fig. 3a-b**).

**Figure 2.**
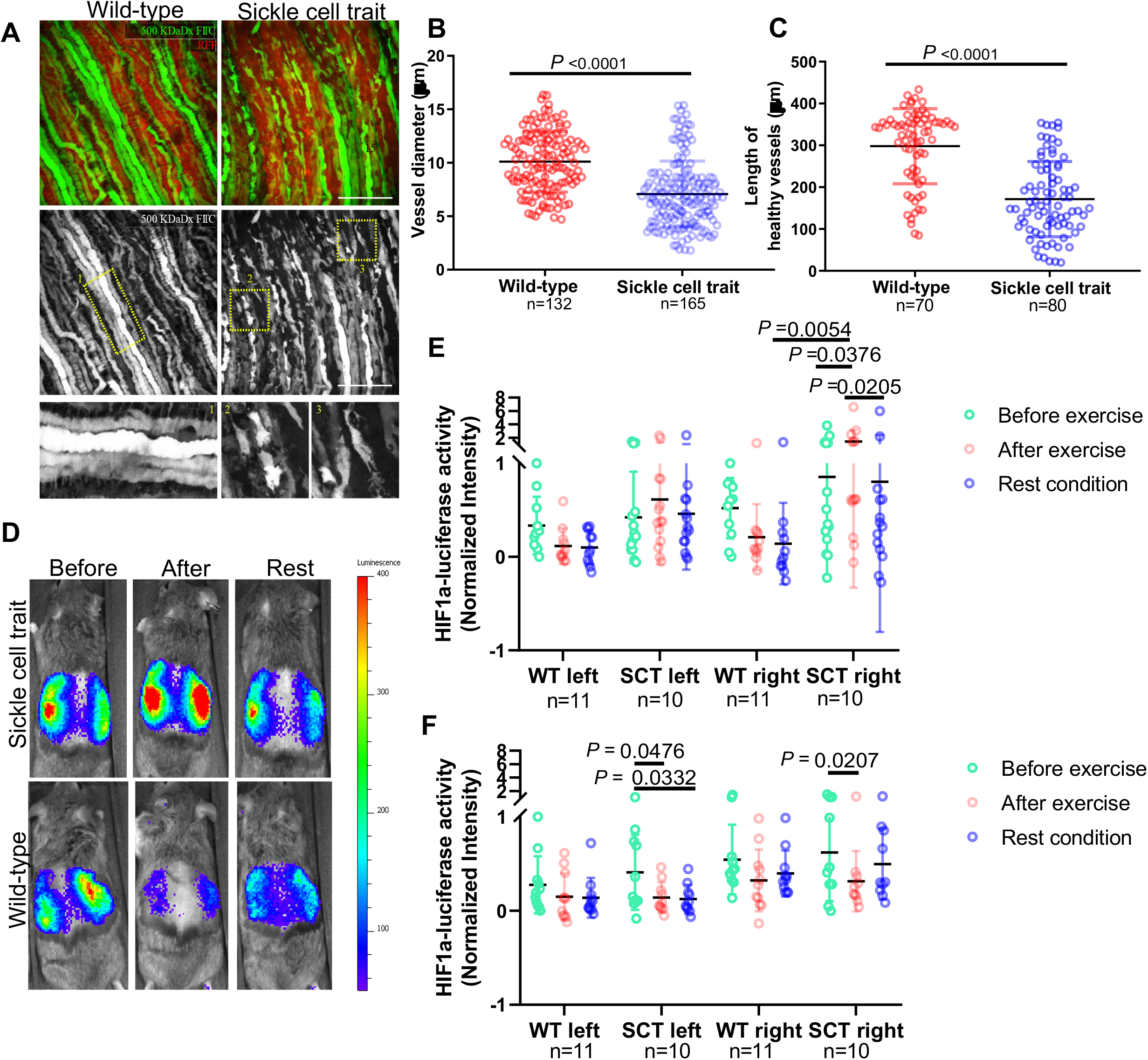
**(A)** 3D image reconstruction of renal epithelia (RFP) and FITC-dextran (GFP) in adult mice (n=4-5) with kidney-specific *CDH16*^*Cre*^ and conditional *R26*^*LSL-Tom*^ shows hypoperfusion in the inner medulla of mice with SCT. **(B, C)** Quantification of the diameter **(B)** and length **(C)** of the renal blood vessels.(10 vessels/image, 3 locations/vessel). *P* values were calculated by student’s t test (average of single values/mouse). **(D)** IVIS imaging of wild-type (n=11) and SCT (n=10) mice harboring the HIF1α oxygen-dependent degradation domain fused to firefly luciferase before high-intensity exercise, immediately after, and after one hour of rest. **(E-F)** Intensity of HIF1α-luciferase activity after high-intensity exercise **(E)** and moderate-intensity exercise **(F)**. Two-way ANOVA and Tukey multiple comparison tests were used to calculate the *P* values of the changes in HIF1α–luciferase activity in adult SCT (n=10) and wild-type (n=11) mice after exercise and rest.

Complications related to hypoxia in the setting of high-intensity exercise have previously been reported among athletes and military recruits with SCT.^4,5^ To test whether high-intensity exercise would worsen renal hypoxia among SCT mice, we generated a GEMM of SCT by crossing the hα/hα::β^S^/β^S^ strain with the *Gt(ROSA)26Sor*^*tm2(HIF1A/luc)Kael*^ strain, allowing non-invasive monitoring of HIF1α activity in response to hypoxia using the *in vivo* imaging system (IVIS) (**Supplementary Fig. 2b**). We evaluated wild-type and SCT mice with two treadmill exercise intensities: moderate-intensity exercise (12 meters/minute), which correlates with 65% to 70% VO2 max, and high-intensity exercise (15 meters/minute), which correlates with 80% VO2 max as established in a prior study.^6^ Luciferase fluorescence was measured prior to exercise, immediately after exercise, and after one hour of rest (**Fig. 2d**). High-intensity exercise resulted in significantly higher HIF1α activation among SCT mice compared to wild-type (*P* = 0.0054), which was most pronounced within the right kidney (*P =* 0.0376) (**Fig. 2e**). Conversely, when exposed to moderate-intensity exercise, there was a significant decrease in HIF1α activation in the SCT mice for both the left (*P* = 0.0476) and right *(P =* 0.0207) kidneys. There was no significant change in wild-type mice (**Fig. 2f**). While high-intensity exercise exacerbated renal hypoxia in SCT, moderate-intensity exercise mitigated hypoxia, particularly in SCT mice. The increased tissue hypoxia in the right kidney of SCT mice corresponded to the distinct predilection of RMC to arise from the right kidney in humans. Similar to humans, the right renal artery length in the C57BL/6J mice used in our study was longer than the left (**Supplementary Fig. 2d-e**).

Given the lethality of RMC, it is essential to elucidate potentially modifiable risk factors for disease development. Our study demonstrates that high-but not moderate-intensity exercise is a risk factor for RMC among individuals with SCT and serves as a foundation for future investigations into RMC tumorigenesis. These findings support prior studies recommending low/moderate-intensity exercise training to reduce complications related to sickle hemoglobinopathies^5,7,8^ and should be considered when counseling and monitoring individuals with these conditions, particularly those occupationally engaged in high-intensity exercise.

## METHODS

### RMC patient clinical data

Clinical and pathologic data from pathologically confirmed RMC patients treated at a single institution were collected. All histology slides were reviewed by a genitourinary pathology expert and the RMC samples were all confirmed to be SMARCB1 negative by immunohistochemistry (IHC) using purified mouse anti-BAF47 Clone 25/BAF47 (BD Biosciences). Clinical variables included age, biologic sex, race, presence of sickle hemoglobinopathy, comorbidities, smoking history, body mass index (BMI), family history of RCC, presence of symptomatic disease, clinical TNM stage, and metastatic locations. Patients were also evaluated for reported regular physical activity or military service. Reported involvement in organized athletic activities, exercise ≥ 3 times per week, participation in endurance racing, or prior military service were retrospectively evaluated for each patient and recorded if present. A binary “activity index” was created whereby patients with involvement in any of the above listed exercise activities were coded as “1” (“yes”) and those without involvement in any exercise activity were coded as “0” (“no”).

We generated a matched control group to determine if reported exercise (as measured by the activity index) and anthropometric measurements were associated with RMC using a case-control approach. Because it is well established that advanced malignancies, including advanced RMC, can lead to sarcopenia and cachexia,^9^ the matched control patients (all evaluated in the same department as the patients with RMC) also had stage 3 or 4 genitourinary malignancies and were matched with each RMC patient in a 2:1 ratio whenever possible based on age and biologic sex. The types of genitourinary malignancies included testicular cancer, RCC, prostate cancer, and bladder cancer (**Supplementary table 4**).

### Anthropometric analysis

We objectively measured the cross-sectional area of skeletal muscle (SM) and subcutaneous adipose (SA) compartments using computed tomography (CT) imaging studies performed during the initial staging of each patient’s cancer. Images were analyzed at the initial diagnosis of cancer prior to initiation of systemic therapy to avoid the changes induced by systemic therapy on either skeletal muscle or subcutaneous adipose tissue. We then compared the two compartments between the RMC patients and the matched controls. The SM and SA cross-sectional areas were measured using axial images at the midpoint of the L3 vertebral body as performed in prior studies.^10^ Cross-sectional areas were standardized to the square of height in meters for each patient. Analysis was performed using SliceOMatic v5.0 (TomoVision, Magog, Canada). The density boundaries used to tag and quantify SM were −29 to +150 and the boundaries for SA were −190 to −30. Skeletal muscle was tagged in red and SA was tagged in green.

### Epidemiologic comparison

We hypothesized that active-duty military service may increase the rate of high-intensity exercise and be more prevalent in our RMC cohort. We compared the rate of military service among our RMC cohort to that of a similarly aged (age 20-40 years old) US population of black individuals with SCT. To estimate this population, we used US census bureau data from 2018 which report that 41,617,764 individuals identify as black (about 12.7% of the U.S. population).^11^ Of this population, about 35% (14,566,217) are between 20 and 40 years old, which is the age most likely to be affected by RMC.^11^ Department of defense data from 2018 report that there are 222,705 black individuals serving on active duty age 20 to 40 years old.^12^ Sickle cell trait affects about 8% of black individuals^3^; thus, there are about 1,165,297 black individuals age 20 to 40 years with SCT in the United States, of which a total of 17,816 (1.5%) are on active duty in the US military.

### Prospective exercise evaluation

We prospectively gathered detailed information on the exercise activity of 7 additional patients with RMC seen as new patients at our institution from March 2020 until January 2021. These patients were asked to grade their exercise intensity as high, moderate, or none. High-intensity exercise was defined as achieving near maximal effort by performing at ≥80% maximal heart rate during each exercise session.^13^ Moderate intensity exercise was defined as exercise performed in a continuous manner at lower intensities than high-intensity exercise during each exercise session.^13^ Patients who did not participate in regular exercise activities were reported as “none”. Patients were also asked how much time per week was spent on exercise. Lastly, we recorded the type of exercise activities patients participated in and how long they had been involved in these activities prior to the diagnosis of RMC.

### Mouse strains

The Townes model of SCT (hα/hα::β^A^/β^S^) was generated by Dr. Tim Townes’s laboratory and obtained through Jackson Laboratory (Stock No. 013071).^14^ The *Cdh16*-Cre strain was generated by Peter Igarashi’s laboratory and obtained through Jackson Laboratory (Stock No. 012237). The *Gt(ROSA)26Sor*^*tm2(HIF1A/luc)Kael*^ strain was generated by Dr. William G. Kaelin’s laboratory and obtained through Jackson Laboratory (Stock No. 006206).^15^ The *Rosa26*^*LSL-TdT*^ was generated by Dr. Hongkui Zeng’s laboratory and obtained through the Jackson Laboratory (Stock No: 007908).^16^ Strains were kept in a mixed C57BL/6J and 129Sv/Jae background. All animal studies and procedures were approved by the UTMDACC Institutional Animal Care and Use Committee.

### Genotyping conditions

#### Townes model (hα/hα::β^A^/β^S^)

The common (mouse beta KI) forward primer is 5’-TTGAGCAATGTG GACAGAGAAGG; the Beta A reverse primer is 5’-GTTTAGCCAGGGACCGTTTCAG; the Beta S reverse primer is 5’-AATTCTGGCTTATCGGAGGCAAG; the reverse primer for mouse beta wild type is 5’-ATGTCAGAAGCAAATGTGAGGAGCA. The separated PCR conditions are as follows: 94°C for 2 minutes, continue PCR for 10 cycles at 94°C for 20 seconds, 65°C with 0.5°C decrease per cycle for 15 seconds, 68°C for 10 seconds, continue PCR for 28 cycles at 94°C for 15 seconds, 60°C for 15 seconds, 72°C for 10 seconds.

#### *Cdh16*-Cre

The forward primer for the transgene is 5’-GCAGATCTGGCTCTCCAAAG; the forward primer for the internal positive control is 5’-CAAATGTTGCTTGTCTGGTG; the reverse primer for the internal positive control is 5’-GTCAGTCGAGTGCACAGTTT; the reverse primer for the transgene is 5’-AGGCAAATTTTGGTGTACGG. The touchdown PCR cycling conditions are as follows: 94°C for 2 minutes, continue PCR for 10 cycles at 94°C for 20 seconds, 65°C with 0.5°C decrease per cycle for 15 seconds, 68°C for 10 seconds, continue PCR for 28 cycles at 94°C for 15 seconds, 60°C for 15 seconds, 72°C for 10 seconds.

#### Gt(ROSA)26Sor^tm2(HIF1A/luc)Kael^

The mutant forward primer is 5’-CGGTATCGTAGAGTCGAGGCC; the mutant reverse primer is 5’-GGTAGTGGTGGCATTAGCAGTAG; the wild-type forward primer is 5’-AAGGGAGCTGCAGTGGAGTA; the wild-type reverse primer is 5’-CCGAAAATCTGTGGGAAGTC. The separated PCR conditions are as follows: 94°C for 2 minutes, continue PCR for 10 cycles at 94°C for 20 seconds, 65°C with 0.5°C decrease per cycle for 15 seconds, 68°C for 10 seconds, continue PCR for 28 cycles at 94°C for 15 seconds, 60°C for 15 seconds, 72°C for 10 seconds.

#### Rosa26^LSL-TdT^ allele

The forward primer for wild-type Tomato is 5’-AAGGGAGCTGCAGTGGAGTA; the reverse primer for wild-type Tomato is 5’-CCGAAAAATCTGTGGGAAGTC; the forward primer for Tomato mutant is 5’-CTGTTCCTGTACGGCATGG; the reverse primer for Tomato mutant is 5’-GGCATTAAAGCAGCGTATC. The touchdown PCR cycling conditions are as follows: 10 cycles at 94°C for 20 seconds, 65°C and 0.5°C per cycle decrease for 15 seconds, 68°C for 10 seconds. Continue the PCR reaction for 28 cycles at 94°C for 15 seconds, 60°C for 15 seconds, 72°C for 10 seconds.

### IVIS imaging

*In vivo* imaging system (IVIS) was used to detect luciferase activity in mice. Mice with pigmented fur were shaved prior to imaging to prevent background caused by melanin in fur. Wild-type mice and mice with SCT were injected with 100 µL of 150 mg/kg of d-luciferin bioluminescence substrate (Perkin Elmer) via intraperitoneal injection (IP) and imaged five minutes after the injection. Living Image 4.3 software was used for analyzing images after image acquisition.

### Forced exercise

Adult untrained mice, n=11 wild-type mice and n=10 SCT mice, were placed on a six-lane mechanical treadmill (Columbus Instruments) that was quickly brought up 12 to 15 meters/minute to model acute exercise. Each session lasted 10 minutes. Mice were encouraged to continue running by using a soft-bristled tube cleaning brush to gently push them from behind if they began to lag. For IVIS imaging studies, mice were imaged prior to exercise, after exercise, and one hour after exercise. Moderate-intensity exercise was defined as 12 meters/minute, which correlates with 65% to 75% VO2 max in humans, i.e. a brisk walk.^6^ High-intensity exercise was defined as 15 meters/minute, which correlates with approximately 80% VO2 max in humans.^6^Luciferase activity was normalized to the left wild-type kidney per mouse for further statistical analysis.

### Ex vivo multiphoton microscopy analysis

Four adult wild-type mice and five SCT mice were injected with fluorescein isothiocyanate (FITC)–dextran 500,000 KDa (Sigma Aldrich) and euthanized after five minutes. The large molecular weight dextran was used to prevent filtration of dextran outside the renal tubules. Kidneys were removed and monitored by multiphoton microscopy (Trimscope II, LaVision BioTech) using a long working distance 25x NA 1.05 oil/water objective (Olympus). The microscope was equipped with three titanium (Ti):sapphire lasers (Chameleon-XR, Coherent) and two optical parametric oscillators (APE/Coherent), resulting in a tunable excitation range from 800 to 1300 nm.^17^ Detection of blood vessels and kidney cells was achieved by spectral separation of FITC (525/50, 920 nm) and RFP (595/40, 1090 nm), and 3D stacks were obtained for up to 300 µm depth at 10 µm step size. Images from individual 3D stacks were reconstructed and analyzed using FIJI.^18^ Vessel diameter and length were measured manually. The diameter was measured in 10 vessels/image, in three different location within each vessel. The length was measured in 10 vessels/image.

### Pimonidazole hydrochloride (Hypoxyprobe)

Renal hypoxia was measured using pimonidazole hydrochloride (Hypoxyprobe). Adult wild-type mice (n=52) and mice with SCT (n=31) were injected with 100 µL of 60 mg/kg of pimonidazole hydrochloride three hours prior to sacrifice. Mice were then anesthetized using isoflurane (Henry Schein Animal Health), and kidneys were removed while mice were still living in order to prevent background hypoxia signal that can be caused by euthanasia using CO2 asphyxiation. Mice were sacrificed via exsanguination. A scalpel was then used to prepare sagittal kidney sections, which were immediately fixed with 10% formalin for 24 hours at room temperature for further IHC studies. The amount of time between removal of the kidneys from mice and fixing in formalin was minimized in order to accurately measure tissue hypoxia.

### Immunohistochemistry

IHC was performed on FFPE whole kidney sagittal sections. The sagittal kidney sections were fixed in 10% formalin, embedded, and 5 µm sections were cut using a microtome (Leica RM2235). The sections were then baked on slides, de-paraffinized, and treated with citrate buffer (Electron Microscopy Sciences) for antigen retrieval according to the manufacturer’s instructions. Endogenous peroxidases were then inactivated using 3% hydrogen peroxide (Sigma-Aldrich) for 10 minutes followed by washing with phosphate buffer saline. Non-specific signals were then blocked for 20 minutes using Rodent Block M (Biocare Medical). Samples were then stained with 1:200 primary Hypoxyprobe antibody conjugated with horseradish peroxidase (HRP, rabbit, Hypoxyprobe) for 12 hours overnight in 4°C. After an overnight incubation, slides were washed three times using phosphate buffer saline before staining with Rabbit-on-Rodent HRP polymer (Biocare Medical) for one hour at room temperature. NovaRED peroxidase substrate (Vector Lab) was used for HRP detection (10 minutes of exposure). Hematoxylin was used for counterstaining and eosin was used to detect the cytoplasm of cells in tissue slides. A Nikon EclipseTi microscope and Nikon DS-Fi1 digital camera were used to capture 20x images.

Equal areas of each 20x image were then quantified using ImageJ/FIJI. Using FIJI, the IHC images were deconvoluted into three channels. The optical densities of the DAB channel images were then quantified and graphed into GraphPad for further statistical analysis.

### Statistical analysis

To reduce bias introduced by inappropriate regression adjustment for colliders or mediators rather than confounders, we generated directed acyclic graphs (DAGs) to codify our pre-specified model that guided our statistical analyses evaluating the relationship between reported exercise history, anthropometric measurements, and the diagnosis of RMC.^19^ Multivariable logistic regression models were used to determine the association between reported exercise history or anthropometric measures with RMC diagnosis (compared with diagnosis of the matched control genitourinary malignancies) following adjustment for confounders (age, biologic sex, and race) identified by the DAGs shown in **Supplementary Figure 1**. Overall survival was estimated using the Kaplan-Meier method. Continuous variables were compared between RMC and matched control patients using Wilcoxon rank-sum tests, and categorical variables were compared using Fisher’s exact tests. Analysis was performed using Stata/SE v16.1 (StataCorp, College Station, TX). DAG figures were generated using the R package “dagitty.”^20^ Data were graphed using GraphPad. Two-way ANOVA and Tukey multiple comparison tests were used to compare the changes in HIF1α–luciferase activity in SCT (n=10) and wild-type (n=11) mice after exercise and rest shown in **Fig. 2e-f**.

## Data Availability

Data may be shared upon reasonable request to the corresponding author

## Acknowledgements

Supported in part by the Cancer Center Support Grant to MDACC (grant P30 CA016672) from the National Cancer Institute of the National Institutes of Health and by a Concept award (W81XWH1810570) from the United States Department of Defense.

## Author Contributions

This study was conceptualized by D.D.S., M.S., G.G., and P.M. Methodology was determined by D.D.S., M.S., L.P., E.D., G.G., and P.M. Writing including the original draft, review, and editing was performed by D.D.S., M.S., G.G., and P.M. Writing including review and editing was performed by D.N.T., J.B., R.X., M.W.S., M.L.V., P.R., M.H.G.K., N.H.P., A.Y.S., A.C., T.P.H., K.L.S., C.L., C.L.W., C.G.W., J.A.K., G.F.D., and N.M.T. The investigation, visualization, and formal analyses were performed by D.D.S., M.S., L.P., G.G., and P.M. Resources were provided by E.D., D.S.S., G.G., P.M., M.W.S., M.H.G.K., N.H.P., C.L.W., C.G.W., J.A.K., and N.M.T. This work was supervised by N.M.T., G.G., and P.M.

---

**Supplementary Figure 1.**
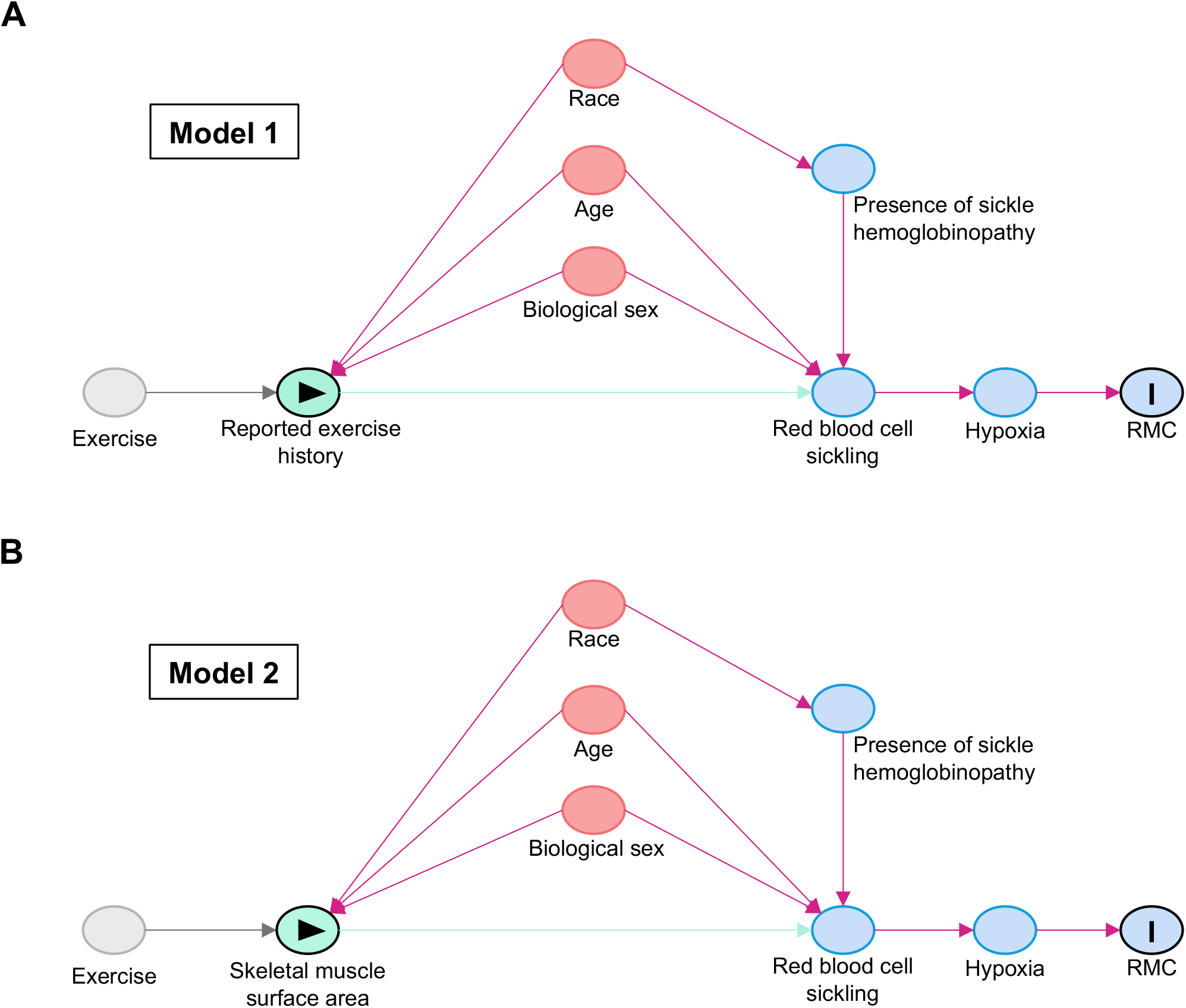
Directed acyclic graphs. Directed acyclic graphs (DAGs) model the causal relationships between exposures and outcomes of interest. Arrows indicate a causal interaction between two variables. Green circles with triangle represent the exposure of interest and blue circles with I represent the outcome of interest (development of RMC). Red circles represent confounding variables that should be adjusted for in order to determine the causal effect of the exposure on the outcome. Each model was used to construct a multivariable logistic regression adjusting for confounding variables identified in the DAG. **(A)** Model 1 demonstrates the causal relationship between reported exercise history and development of RMC. **(B)** Model 2 demonstrates the causal relationship between standardized skeletal muscle surface area and development of RMC.

**Supplementary Figure 2.**
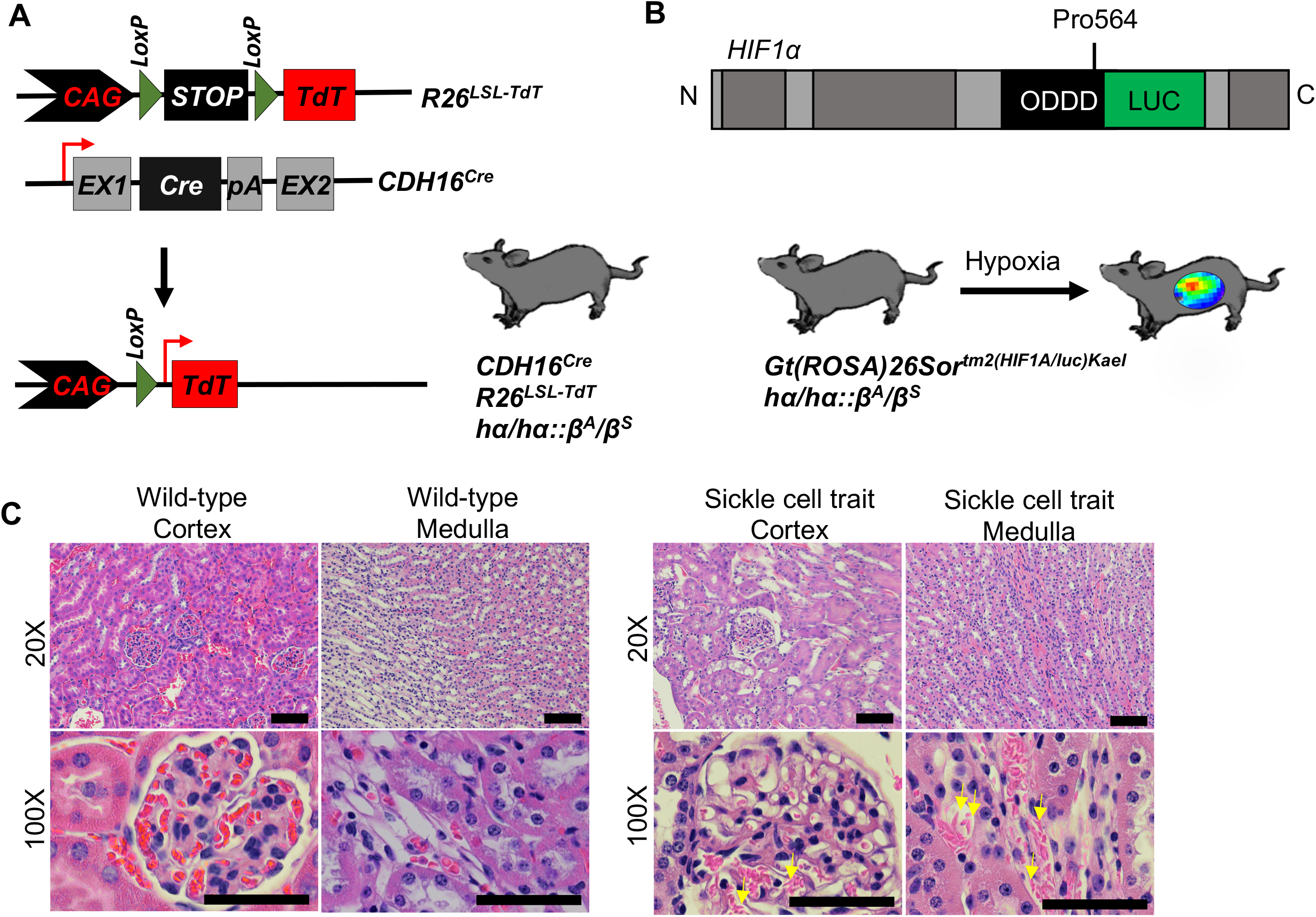
Mouse models of sickle cell trait. **(A)** Schematic of genetically engineered mouse model (GEMM) of SCT. The *CDH16*^*Cre*^ strain was crossed with the conditional *Rosa26*^*LSL-TdT*^ strain and the hα/hα::β^S^/β^S^ strain to generate a GEMM of SCT (hα/hα::β^A^/β^S^) that allows for tissue specific activation of TdTomato (TdT) fluorescence reporter in the kidney epithelium. **(B)** Schematic of GEMM of SCT with HIF1α oxygen-dependent degradation domain fused to firefly luciferase, allowing non-invasive monitoring of HIF1α activity in response to hypoxia using the *in vivo* imaging system (IVIS). The hα/hα::β^S^/β^S^ strain was crossed with the *Gt(ROSA)26Sor*^*tm2(HIF1A/luc)Kael*^ strain. **(C)** Hematoxylin and eosin staining of kidney tissue from wild-type mice (left) and mice with SCT (right). Sickle morphology is apparent in mice with SCT. Sickled RBCs have spindle-like morphology and are indicated with yellow arrows.

**Supplementary Figure 3.**
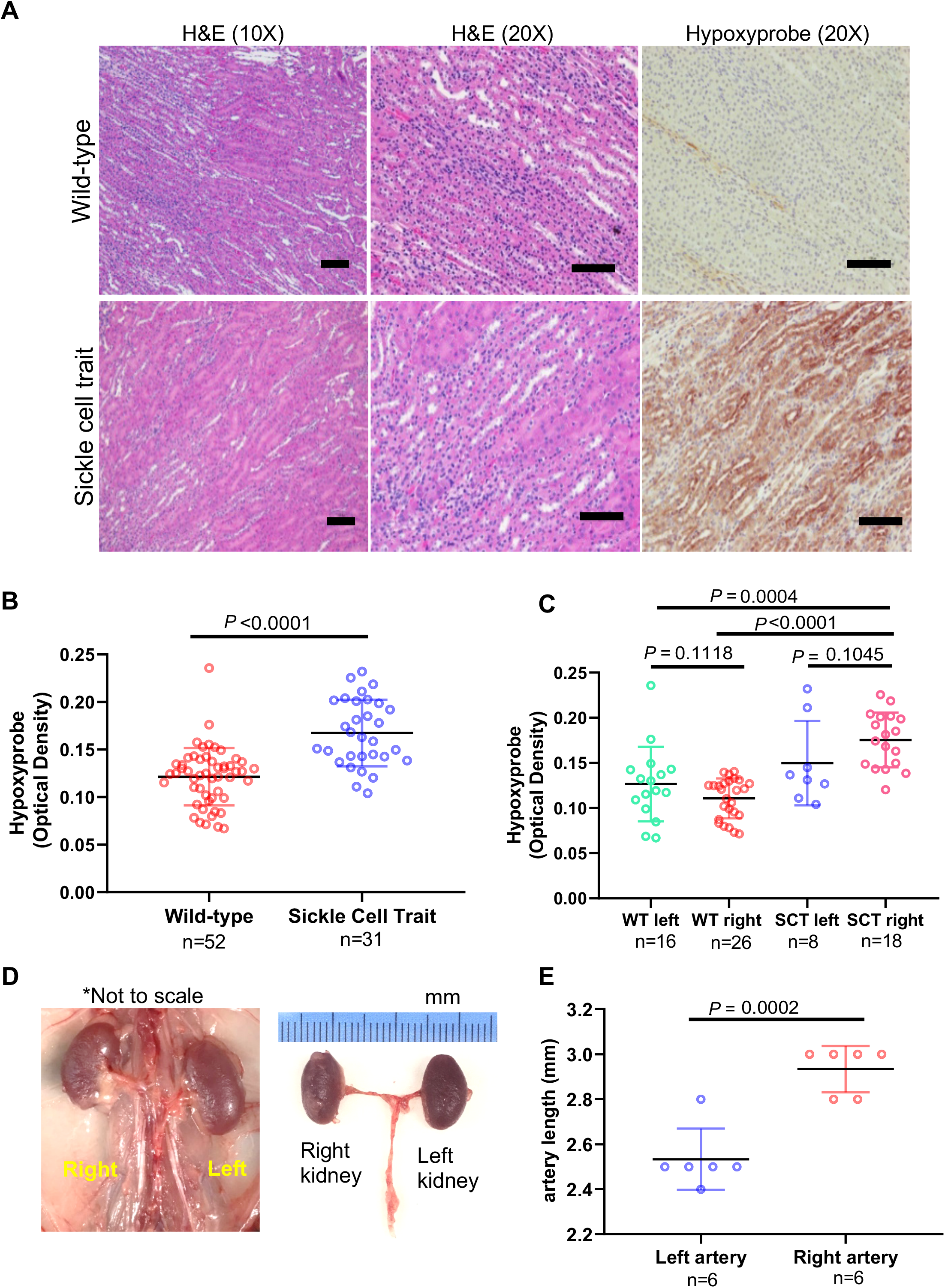
Quantification of renal hypoxia and mouse kidney anatomy. **(A)** Hypoxyprobe (pimonidazole hydrochloride) was injected in the intraperitoneal cavity of GEMM of SCT with kidney-specific *CDH16*^*Cre*^ and conditional *R26*^*LSL-Tom*^ three hours prior to euthanasia and harvesting of kidneys. Kidneys were processed and immunohistochemistry with the Hypoxyprobe antibody was used to show hypoxia in the inner medulla of wild-type mice (top) and mice with SCT (bottom). **(B-C)** Quantification of the optical density of horseradish peroxidase staining for 20x images was done using ImageJ. Comparisons were made between wild-type (n=52) and SCT mice (n=31) **(B)**. Additionally, quantification was stratified by kidney laterality within SCT (left kidneys, n=8; right kidneys, n=18) and wild-type (left kidneys, n=16; right kidneys, n=26) mice **(C)**. Data are expressed as mean value ± SD, with *P* value calculated by student’s *t* test. **(D)** Gross anatomy of C57BL/6J mouse showing difference in the lengths between the right and left renal arteries. **(E)** Quantified difference in the length of left and right C57BL/6J mouse renal arteries from six adult mice. Data are expressed as mean value ± SD, with *P* value calculated by student’s *t* test.

## TABLES

**Supplementary Table 1.** Clinical characteristics of RMC patients and matched control patients.

| Variable | RMC (n=71) | Matched (n=122) | P value |
| --- | --- | --- | --- |
| Median age (IQR) | 29.6 (23.2-37.5) | 29.9 (23.8-39.6) | 0.5 |
| Biological sex, no. (%) |  |  | 0.7 |
| Male | 49 (69) | 87 (71.3) |  |
| Female | 22 (31.0) | 35 (28.7) |  |
| Race, no. (%) |  |  | <0.001 |
| African American | 61 (85.9) | 47 (38.5) |  |
| White | 7 (9.9) | 66 (54.1) |  |
| Hispanic | 2 (2.8) | 4 (3.3) |  |
| Asian | 1 (1.4) | 5 (4.1) |  |
| Sickle hemoglobinopathy, no. (%) | 71 (100) | 0 | <0.001 |
| Sickle cell trait | 70 (98.6) |  |  |
| Sickle beta-thalassemia | 1 (1.4) |  |  |
| Comorbidities, no. (%) |  |  |  |
| HTN | 12 (16.9) | 33 (27.1) | 0.2 |
| Other comorbidities | 8 (11.3) | 58 (47.5) | <0.001 |
| Median BMI, (IQR) | 27 (24.3-29.9) | 27.1 (23.9-31.6) | 0.7 |
| Smoking history, no. (%) | 15 (21.1) | 56 (45.9) | 0.001 |
| Exercise history, no. (%) | 36 (50.7) | 18 (14.8) | <0.001 |
| Athlete, no. (%) | 24 (33.8) | 5 (4.1) | <0.001 |
| Military, no. (%) | 7 (9.9) | 4 (3.3) | 0.005 |
| Activity index, no. (%) | 47 (66.2) | 20 (16.4) | <0.001 |

**Supplementary Table 2.** Additional clinical characteristics of RMC patients.

| **Variable** | **RMC (n=71)** |
| --- | --- |
| Family history RCC, no. (%) | 4 (5.6) |
| Symptoms |  |
| Pain | 55 (77.5) |
| Hematuria | 39 (54.9) |
| Weight loss | 19 (26.8) |
| Fever | 1 (1.4) |
| Chills | 1 (1.4) |
| Night sweats | 3 (4.2) |
| Localized at diagnosis | 8 (11.3) |
| Metastatic at diagnosis | 63 (88.7) |
| T stage |  |
| T1-T2 | 66 (93.0) |
| T3-T4 | 3 (4.2) |
| Unknown | 2 (2.8) |
| Max tumor diameter, median (IQR) | 5.8cm (5-7cm) |
| RMC laterality |  |
| Right kidney | 54 (76.1) |
| Left kidney | 17 (23.9) |
| Multiple metastatic locations | 55 (77.5) |
| Nephrectomy | 50 (70.4) |

**Supplementary Table 3.** Metastatic locations among RMC patients with metastatic disease at initial diagnosis.

| **Metastatic site** | **N=63 (%)** |
| --- | --- |
| Retroperitoneal nodes | 52 (82.5) |
| Lungs | 45 (71.4) |
| Bone | 15 (23.8) |
| Liver | 14 (22.2) |
| Hilar LN | 13 (20.6) |
| Soft tissue | 5 (7.9) |
| Supraclavicular LN | 4 (6.4) |
| Ipsilateral adrenal | 4 (6.4) |
| Pancreas | 2 (3.2) |
| Thyroid | 2 (3.2) |
| Mesentery | 1 (1.6) |
| Ovary | 1 (1.6) |
| Spleen | 1 (1.6) |
| Brain | 1 (1.6) |
| Bowel | 0 |
| Contralateral kidney | 0 |
| Contralateral adrenal | 0 |

**Supplementary Table 4.** Cancer type and stage for matched control patients.

| **Matched Patient**<br>**Cancer Type** | **N = 122 (%)** | **Stage III, n (%)** | **Stage IV, n (%)** |
| --- | --- | --- | --- |
| Testicular cancer | 46 (37.7) | 46 (100) | 0 |
| Renal cell carcinoma | 46 (37.7) | 13 (28.3) | 33 (71.8) |
| Bladder cancer | 21 (17.2) | 4 (19.0) | 17 (81.0) |
| Prostate cancer | 9 (7.4) | 0 | 9 (100) |

**Supplementary table 5.**
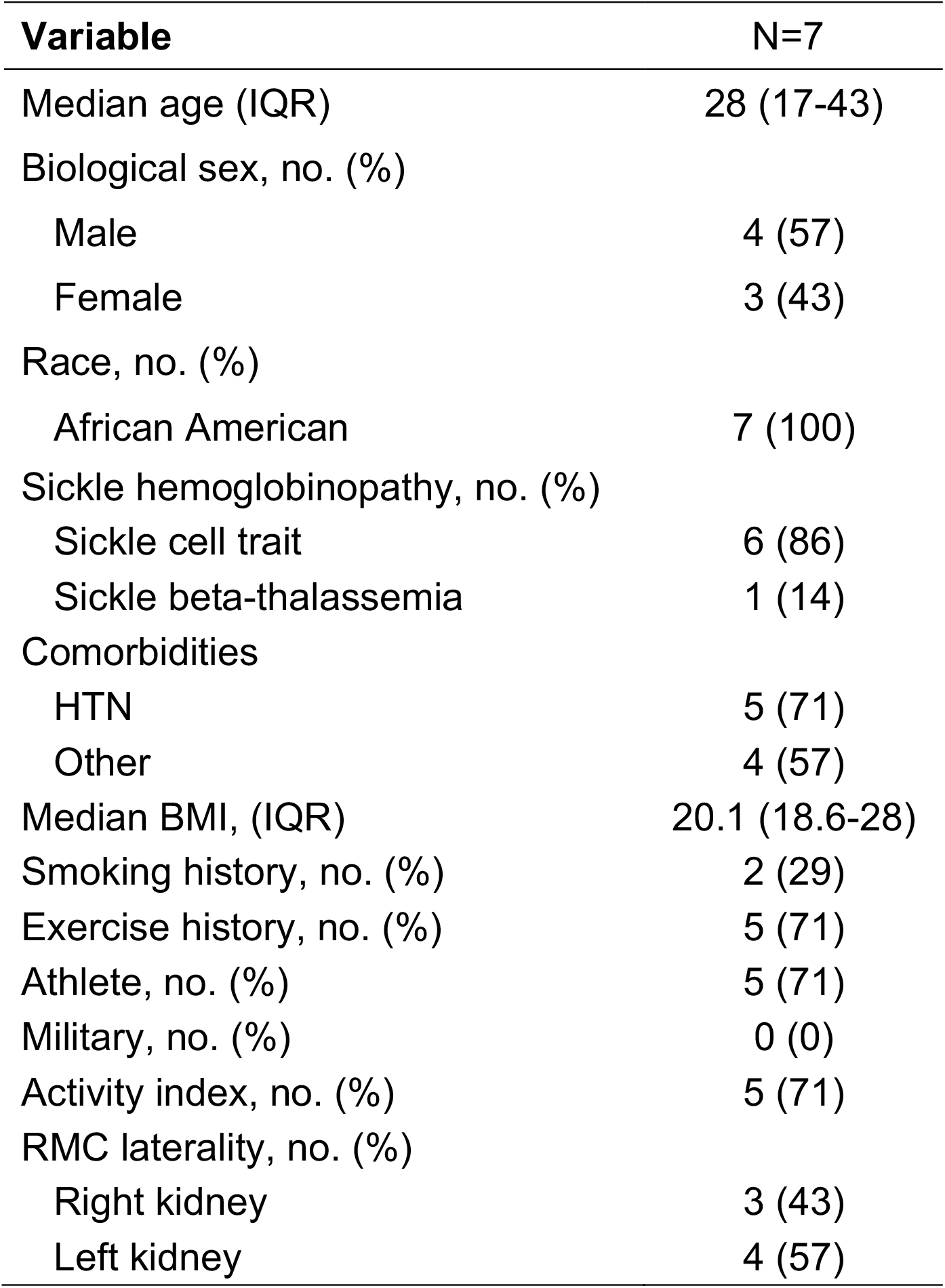
Clinical characteristics of prospectively evaluated patients with RMC.

**Supplementary table 6.** Prospectively collected exercise history of seven patients with RMC.

| ID | Exercise intensity | Exercise time per week | Participation prior to RMC diagnosis | Exercise Description | Primary tumor laterality |
| --- | --- | --- | --- | --- | --- |
| 1 | High | ≥3 hours | >3 years | Softball and swimming | Left |
| 2 | High | ≥2 hours | >3 years | Basketball and frequent gym exercise | Right |
| 3 | High | ≥2 hours | >3 years | Basketball, soccer, weightlifting | Right |
| 4 | High | ≥3 hours | >3 years | Professional athlete | Left |
| 5 | None | None | None | Sedentary lifestyle | Left |
| 6 | High | ≥3 hours | >3 years | Track, martial arts, and frequent gym exercise | Right |
| 7 | None | None | None | Sedentary lifestyle | Left |

